# Age-Dependent Effects of UV Exposure and Xeroderma Pigmentosum Group A on DNA Damage, Repair Mechanisms, Genomic Instability, Cancer Risk, and Neurological Disorders

**DOI:** 10.1101/2024.07.22.24310800

**Authors:** WR Danter

**Affiliations:** 123Genetix Inc

**Keywords:** Xeroderma Pigmentosum Group A (XPA), UV Exposure, DNA Damage Repair, Nucleotide Excision Repair (NER), Genomic Instability, Neurological Disorders, aiHumanoids, Virtual Longitudinal Study, Skin Cancer Risk

## Abstract

**Background:** Xeroderma pigmentosum, complementation group A (XPA), is a rare genetic disorder characterized by marked sensitivity to ultraviolet (UV) radiation, leading to increased risks of skin cancer, accelerated aging, and significant neurologic disorders. XPA prominently impacts DNA repair mechanisms, specifically nucleotide excision repair (NER), which is crucial for correcting UV-induced DNA damage.

**Methods:** This study utilized an advanced aiHumanoids platform to simulate the disease progression in individuals with XPA from birth to age 20 years. The virtual longitudinal study assessed the impacts of moderate and severe XPA under various UV exposure scenarios. The research included 25 age-matched wild-type controls to elucidate the comparative effects of XPA on DNA damage, genomic instability, cancer risk, and neurological outcomes.

**Results:** Using Wilcoxon sign rank p values and Cliff’s delta estimates of true effect size, the aiHumanoid simulations revealed significant differences in DNA repair efficiency between XPA affected and control groups, with pronounced deficits in XPA cohorts under UV exposure. Genomic instability and skin cancer risks were consistently elevated across all XPA simulations, particularly under UV stress. Neurological assessments indicated greater susceptibility to disorders in younger XPA subjects, with effects moderating somewhat with age.

**Conclusion:** The aiHumanoid platform provided novel insights into the progression of XPA, highlighting the severe impact of UV exposure on individuals with this condition. These findings advocate for early intervention strategies and underscore the necessity for rigorous protective measures against UV radiation, especially in younger populations. This research contributes to our further understanding of XPA, potentially guiding future therapeutic developments including early stage virtual drug trials and preventive approaches personalized to individual risk profiles.

## INTRODUCTION

Xeroderma pigmentosum, complementation group A (XPA), is a rare, debilitating genetic disorder characterized by extreme sensitivity to ultraviolet (UV) radiation, leading to a high risk of skin cancers and accelerated skin aging [1]. Among the xeroderma pigmentosum (XP) subtypes, XPA is one of the more severe forms due to a marked deficiency in nucleotide excision repair (NER), a critical DNA repair mechanism [2,3]. The global incidence of XPA, which varies widely, is approximately 1 in 1,000,000 individuals, making it a rare but profoundly impactful condition [4]. Patients with XPA not only face increased adverse skin effects but are also at an increased risk of developing neurological disorders, a problem that is less common in some other XP subtypes [5]. For example, XP subtypes A, D, and G are more commonly associated with neurological disorders, while the B, C, E, F, and V forms primarily affect the skin [6,7].

Traditionally, longitudinal studies that track the progression of genetic disorders like XPA face significant hurdles, including (i) the rarity of the condition, (ii) the extensive duration needed to acquire reliable data, and (iii) logistical challenges in managing conditions with consistent levels of UV exposure [8]. These challenges necessitate innovative approaches, including AI, to study disease progression and evaluate potential treatments over a subject’s lifetime [9].

The current virtual longitudinal study uses a novel aiHumanoids platform to simulate the early life course of individuals with XPA from birth to age 20 years. Our study uniquely compares moderate and severe forms of XPA under controlled, moderate UV exposure scenarios as well as in the absence of UV exposure, using a cohort of 25 age-matched wild-type (WT) controls for baseline comparisons. By employing advanced computational modeling and aiHumanoid simulations, this approach allows for a detailed, dynamic exploration of the disease’s progression, both dermatological and neurological, and the impact of environmental factors like UV exposure on disease outcomes [10,11].

Our aiHumanoid platform overcomes the logistical limitations of traditional longitudinal studies while providing a controlled environment to systematically assess the long-term effectiveness of therapeutic strategies and the critical interactions between genetic predisposition and environmental factors like UV exposure [12]. Our findings aim to offer unique new insights into the early onset and progression of XPA, facilitating the development of targeted therapies via virtual early stage drug trials [13,14] and preventive measures tailored to individual risk profiles, with particular attention to mitigating the significant neurological disorders seen in many of these patients [5].

## METHODS

### Overview

The current study utilizes aiHumanoid simulations as virtual subjects in a longitudinal investigation into the development and progression of XPA from birth to age 20 years. We aim to better understand the systemic impact of an XPA mutation with and without exposure to moderate UV radiation. By moderate UV exposure we mean a simulated level of 5.5 out of 11+ on the UV index. The absence of UV exposure was simulated by setting the UV index value at 1.

In this first phase of our XP project, we will focus on the impact of moderate and severe XPA mutations on cutaneous and nervous system markers and outcomes. We anticipate that future research will utilize a similar approach to develop early stage virtual drug trials aimed at identifying phenotype modifying therapies for XPA and other XP complementation groups.

#### 1. Updating the aiHumanoid Simulation to v8.4

The previous version 8.3 of the aiHumanoid [15,16] underwent revisions to v8.4.2. The main differences are that the revised version integrates updated simulations for specific XP associated mutations and an updated subsystem for the diagnosis of XPA in children and adolescents. As before, the number of integrated organoid simulations remains at 21. The literature validation of the WT and XPA aiHumanoid simulations employed the same approach used in previous versions to create the updated simulations comprising v8.4.2.

#### 2. XPA Validation Profile in the aiHumanoid Simulations

To confirm a diagnosis of XPA in the affected aiHumanoid simulated young subjects, a list of nineteen genotypic and phenotypic features was assembled from the literature for evaluation and are presented in Appendix A. All features were statistically significantly different from controls for multiple age matched cohorts and regarding the diagnosis of XPA. The present analysis employed a combination of the nonparametric Wilcoxon signed rank test and the Cliff’s delta effect size estimates.

#### 3. Study Design and Objectives

This project is our most recent virtual longitudinal study using the aiHumanoid simulations. The objectives of this study were: (i) to evaluate the impact of moderate UV exposure on young subjects with Moderate XPA or Severe XPA compared to WT/Healthy subjects, (ii) to better understand XPA development and progression from birth to age 20 years, and (iii) to evaluate a panel of disease features for the purpose of conducting virtual drug trials to identify potential phenotype modifying therapies.

### The Virtual subjects used in this study

The profiles for twenty-five unique and healthy virtual young subjects were synthesized by GPT-4, an advanced large language model (December 2023 version,) at https://chat.openai.com/). GPT4 used its extensive database encompassing medical literature, patient profiles, and related clinical information, to synthesize diverse and representative draft profiles for twenty-five healthy children. Each subject profile was reviewed by an experienced physician prior to enrolment. This longitudinal study design permitted us to create a virtual study with 7 genotypic cohorts (see Table 1) X 6 (age groups) X 25 (virtual subjects), the equivalent of data from 1050 young subjects. The virtual subjects used in this study serve as hypothetical, but commonly encountered population based examples of risks associated with the development of XPA in children in specific affected cohorts but do not represent actual individuals or precise medical histories.

**Table 1:**
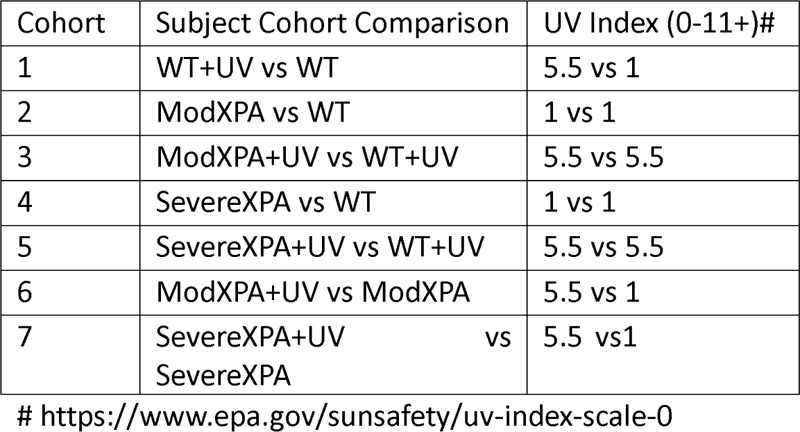
Summary of Cohorts, Comparisons, and UV Index values for this virtual longitudinal study of XPA.

Inclusion Criteria (WT/Healthy cohorts):

1. Generally healthy at birth
2. Ages Birth (0 years) to 20 years
3. Approximately equal representation of boys and girls
4. All required individual data are available

Exclusion criteria:

1. Age greater than 20 years of age
2. Any documented preexisting genetic abnormalities
3. Any documented disease processes prior to or at birth

### The Affected Subjects

The XPA states examined included: (i) the WT state with and without moderate UV exposure, (ii) moderate XPA with and without UV exposure and (iii) severe XPA with and without UV exposure. The six age cohorts of twenty-five healthy subjects each underwent AI gene editing to introduce loss of function (LOF) mutations for each of the six XPA associated cohorts studied [17]. This process created forty two highly matched cohorts where the only difference was the presence or absence of a specific gene mutation. In these well-matched cohorts, properties like obesity, hypertension and Type 2 Diabetes are emergent properties primarily associated with age. The virtual approach has the major advantage that all subjects’ data were available for analysis since there was no attrition which would be common in traditional longitudinal studies of this kind. The data from all cohorts were evaluated beginning at birth (0 years) and continuing at 5-year intervals up to and including age 20 years of age (6 age cohorts).

The distinction between Moderate and Severe XPA was based on estimates of residual normal functioning XPA protein. For the Moderate XPA case an estimate of 25-30% residual protein function was used and for the Severe XPA condition the estimated residual normal protein was <5% [18].

### Statistical Analysis

The Null hypothesis states that there are no statistically significant differences or at least medium effect sizes for the six affected groups compared to the age and UV exposure matched wild type (WT) subjects. The data was not normally distributed, so the non-parametric Wilcoxon signed rank test was used. Given that multiple tests (N=19) were conducted, the conservative Bonferroni correction was applied. The corrected p value to achieve significance therefore became 0.05/19, or < 0.0026 for this study.

The alternative hypothesis states that there are significant differences in the three affected groups compared to the healthy controls (WT).

The true effect size was estimated using Cliff’s delta. Cliff’s delta was used because the data were not normally distributed with a sample size of twenty-five subjects per cohort. To calculate Cliff’s delta the continuous data was transformed into interval data based on whether the data from the affected group was larger or smaller than the unaffected group. The Cliff’s delta was then calculated as (N (larger than) – N (smaller than))/the standard deviation of the differences between the groups (19,20). This produced a range of effect size estimates between -1 (a large negative effect) and +1 (a large positive effect). A value close to zero was interpreted as having no effect. To determine the size of the effect we used the following heuristic scale: d <0.147 (negligible), d = 0.147 to <0.330 (small), d = 0.330 to <0.474 (medium) and d >= 0.474 (large) as suggested in [21]. To compensate for the modest sample size per cohort, all Cliff’s d values were modified using the Hedges correction [22] which was calculated to be 0.984. Final effect size estimates were obtained by multiplying the initial effect sizes by 0.984. The 95% CI around the effect size estimate was calculated using the Standard Error (SE) of the differences/square root of the sample size (2N data points).

## RESULTS

### (1) Wild Type (WT) subjects versus WT subjects plus moderate UV exposure

See Appendix B: Heat Map 1

#### DNA Damage and Repair Pathways (all corrected p values are <0.0026)

Moderate UV exposure resulted in a consistently large effect size (0.984) for DNA damage (CPD/6-4 PPs), DNA NER-GG, DNA NER-TC, DNA replication stress, and ERCC2/XPD across all age groups from newborn (0y) to young adult (20y). Notably, the DNA NER Core exhibited a substantial reduction in effect size at 10 years (0.276), suggesting a temporary decrease in repair efficiency during pre-adolescence.

#### Genome Instability and Cancer Risk (all corrected p values are <0.0026)

Genome instability (CIN) and skin cancer risk (both melanoma and non-melanoma) showed consistently large and effect sizes (0.984) across all age groups, highlighting the persistent risk associated with moderate UV exposure.

#### Neurological Outcomes (corrected p values for Neurodegeneration and Neuroinflammation were not statistically significant (p value ∼1.0) while all other p values were significant at p<0.0026)

While Neuroapoptosis experienced large effects sizes (0.984) across all age groups, Neurodegeneration, and Neuroinflammation displayed negligible effect sizes across all age groups, suggesting no significant impact on these 2 important factors from moderate UV exposure. However, neurodevelopmental disorder (NDD) showed a large negative effect size in younger age groups (-0.984 in 0y to 5y), which decreased in older ages (-0.669 at 15y and 20y), indicating some developmental sensitivity to UV-induced DNA damage. Neurological symptoms and neuronal oxidative stress consistently exhibited large effect sizes (0.984), underscoring the persistent neurological impact of UV exposure.

Skin Aging and Pigmentation (corrected p values are consistently significant for Skin aging and pigmentation at p,0.0026. UV photosensitivity was only significantly different at ages 2-3 years and again at age 20 years)

Skin aging and pigmentation effects were consistently large (0.984) across all ages. UV photosensitivity showed moderate to large negative effect sizes in early childhood (-0.905 at 2-3y and -0.433 at 5y), which became negligible at -0.039 by young adulthood (20y), indicating that younger individuals are more sensitive to UV exposure. By age 20 years the effect size becomes large and more negative at - 0.512.

#### XPA Protein Expression (all corrected p values are <0.0026)

XPA protein expression was high in early years (0y to 5y with a large effect size of 0.984), decreased to moderate at 10 years (0.433), before becoming large again at 15 and 20 years (0.748), consistent with a degree of age-related variability in the normal response to moderate UV exposure.

#### Summary

The data for WT subjects vs WT subjects exposed to moderate UV radiation indicate robust DNA repair mechanisms across all ages for healthy unaffected subjects, with a notable temporary reduction in efficiency during pre-adolescence. The persistent large impact on skin-related outcomes and the developmental sensitivity in younger individuals highlight the importance of protective measures against UV exposure, particularly for younger children. These findings provide crucial insights into the age-dependent responses of healthy individuals to moderate UV exposure, emphasizing the need for targeted UV protection strategies.

### (2) Wild Type (WT) subjects versus subjects with Moderate XPA in the absence of UV exposure See Appendix B: Heat Map 2

#### DNA Damage and Repair Pathways (all corrected p values are <0.0026)

Without UV exposure, XPA subjects displayed consistently large effect sizes (0.984) for DNA damage (CPD/6-4 PPs) across all age groups, indicating substantial DNA damage. Negative effect sizes (-0.984) for DNA NER Core, DNA NER-GG, and DNA NER-TC across all age groups reflect impaired repair mechanisms in XPA subjects compared to WT. DNA replication stress showed moderate to large positive effect sizes (0.590) for most age groups, increasing to 0.905 at 15y, suggesting increased levels of replication stress in older subjects. ERCC2/XPD displayed large and negative effect size (-0.984) across all ages, consistent with impaired DNA repair in XPA subjects.

#### Genome Instability and Cancer Risk (corrected p values were consistently <0.0026 for genome instability and non-melanoma skin cancer risk but failed to achieve significance for melanoma risk)

Genome instability (CIN) and Non melanoma skin cancer risks exhibited large and stable effect sizes (0.984) across all ages, consistent with marked and persistent genome instability in XPA compared to WT subjects. Effect sizes were small to negligible with slight variations for melanoma (-0.039 to 0.118), from birth to 10 years of age after which the effect sizes become large and negative consistent with a decrease in risk of developing melanoma vs non melanoma skin cancer in older children and adolescent XPA subjects not exposed to moderate UV radiation.

#### Neurological Outcomes (all corrected p values are <0.0026)

Neuroapoptosis and neurodegeneration showed large effect sizes (≥0.787) across all age groups, indicating increased potential for neurological damage in XPA. Neurodevelopmental disorder (NDD) consistently displayed large effect sizes (0.984), reflecting substantial developmental impact in XPA. Neurological symptoms, Neuroinflammation, and Neuronal oxidative stress exhibited large effect sizes (0.787), consistent with significant neurological symptoms, increased inflammation and increased oxidative stress in subjects with moderate XPA.

#### Skin Aging and Pigmentation (all corrected p values are <0.0026)

Skin aging consistently displayed large effect sizes (0.984) across all ages, reflecting significant aging effects. Skin pigmentation generally showed large effect sizes (0.984), with some slight variability (0.905 at 0y and 0.827 at 15y), indicating substantial pigmentation changes. UV photosensitivity also consistently showed large effect sizes (0.984), consistent with significant sensitivity.

#### XPA Protein Expression (all corrected p values are <0.0026)

XPA protein expression displayed large negative effect sizes (-0.984) across all age groups, indicating significantly reduced XPA expression in XPA compared to WT young subjects.

#### Summary

These data reveal substantial differences between WT and XPA subjects even in the absence of UV exposure. XPA subjects exhibit consistent DNA repair deficiencies, increased genome instability, significant neurological impacts, and heightened skin aging and cancer risk. These findings continue to confirm the critical role of XPA in maintaining genomic stability and underscore the potentially severe consequences of its deficiency.

### (3) Wild Type (WT) subjects plus UV exposure versus subjects with moderate XPA plus UV exposure See Appendix B: Heat Map 3

#### DNA Damage and Repair Pathways (all corrected p values are <0.0026)

Moderate UV exposure resulted in consistently large effect sizes (0.984) for DNA damage (CPD/6-4 PPs) across all age groups for XPA subjects, indicating substantial DNA damage. Negative large effect sizes (-0.984) for DNA NER Core, DNA NER-GG, and DNA NER-TC across all age groups reflect deficient repair mechanisms in XPA compared to WT. DNA replication stress showed large effect sizes (0.984) for most age groups, with a slight variation at 10y (0.827) and 15y (0.866), indicating significant replication stress across all age groups. ERCC2/XPD displayed a large negative effect size (-0.984) across all ages, consistent with impaired DNA repair.

#### Genome Instability and Cancer Risk (all corrected p values are <0.0026)

Genome instability (CIN) and Non melanoma skin cancer risks consistently exhibited large and stable effect sizes (0.984) across all ages, consistent with persistent genome instability in XPA compared to WT. The risk of developing melanoma is small before age 5 years after which the effect size becomes large and positive with only minimal variability (≥0.905) in older age groups.

#### Neurological Outcomes (all corrected p values are <0.0026 except for NDD at birth and age 5 years)

Neuroapoptosis, Neurodegeneration, and NDD showed large effect sizes (≥0.787) across all age groups, indicating increased neurological damage in XPA. Neuroinflammation also showed large effect sizes (0.787) across all ages, indicating increased inflammation. Neurological symptoms exhibited large effect sizes (0.984), consistent with the potential for significant neurological symptoms in XPA. The effect size for Neuronal oxidative stress was also large (0.787) across all ages, indicating increased oxidative stress.

#### Skin Aging and Pigmentation (all corrected p values are <0.0026)

Skin aging consistently displayed large effect sizes (0.984) across all ages, reflecting significant aging effects. Skin pigmentation also showed large effect sizes (0.984), with very slight variations (0.827 at 0y), indicating substantial pigmentation changes. UV photosensitivity consistently showed large effect sizes (0.984), indicating marked sensitivity.

#### XPA Protein Expression (all corrected p values are <0.0026)

XPA protein expression displayed large negative effect sizes (-0.984) across all age groups, indicating significantly reduced XPA expression in XPA compared to WT.

#### Summary

The data reveal substantial differences between WT and XPA subjects with moderate UV exposure. XPA subjects exhibit consistent DNA repair deficiencies, increased genome instability, significant neurological impacts, and increased skin aging and cancer risk. These findings highlight the critical role of XPA in maintaining genomic stability and underscore the severe consequences of its deficiency.

### (4) Wild Type (WT) subjects versus subjects with severe XPA without UV exposure See Appendix B: Heat Map 4

#### DNA Damage and Repair Pathways (all corrected p values are <0.0026)

Subjects with severe XPA, compared to WT subjects without UV exposure, showed consistently large effect sizes (0.984) for DNA damage (CPD/6-4 PPs) across all age groups, indicating substantial DNA damage. DNA NER Core, DNA NER-GG, and DNA NER-TC exhibited large negative effect sizes (-0.984) consistently, reflecting significant deficiencies in DNA repair mechanisms in XPA subjects. DNA replication stress also demonstrated large effect sizes (0.984) across most age groups, with minor variations at 10y (0.827) and 15y (0.866), indicating elevated replication stress. ERCC2/XPD presented large negative effect sizes (-0.984) across all ages, consistent with impaired DNA repair.

#### Genome Instability and Cancer Risk (all corrected p values are <0.0026)

Genome instability (CIN) displayed large and consistent effect sizes (0.984) across all age groups, indicating marked genomic instability in XPA subjects. Skin cancer risks (both melanoma and non-melanoma) generally showed large effect sizes (0.984) across all ages for non-melanoma skin cancer risk, and minor variations in melanoma risk (0.512 to 0.984), suggesting overall significant skin cancer risks.

#### Neurological Outcomes (all corrected p values are <0.0026)

Neuroapoptosis and neurodegeneration showed large effect sizes (0.787) across all age groups, indicating increased neurological damage in XPA subjects. Neurodevelopmental disorder (NDD) consistently displayed high effect sizes (0.984), reflecting substantial developmental impacts. Neuroinflammation also showed large effect sizes (0.768) across all ages, indicating elevated inflammation. Neurological symptoms exhibited large effect sizes (0.945), suggesting significant neurological symptoms in XPA. Neuronal oxidative stress showed large effect sizes (0.768) across all ages, consistent with increased oxidative stress.

#### Skin Aging and Pigmentation (all corrected p values are <0.0026)

Skin aging consistently displayed large effect sizes (0.945) across all age groups, reflecting market aging effects. Skin pigmentation generally showed large effect sizes (0.938), with some variations (0.827 at 15y), indicating substantial pigmentation changes. UV photosensitivity consistently showed large effect sizes (0.984), indicating marked sensitivity.

#### XPA Protein Expression (all corrected p values are <0.0026)

XPA protein expression exhibited large negative effect sizes (-0.984) across all age groups, indicating a marked decrease in XPA expression in XPA subjects compared to WT.

#### Summary

The data reveal substantial differences between WT and severe XPA subjects in the absence of UV exposure. XPA subjects exhibit consistent DNA repair deficiencies, increased genomic instability, significant neurological impacts, and heightened skin aging and cancer risk. These findings again underscore the critical role of XPA in maintaining genomic stability and emphasize the severe consequences of its deficiency.

### (5) Wild Type (WT) subjects plus UV exposure versus subjects with severe XPA plus UV exposure See Appendix B: Heat Map 5

#### DNA Damage and Repair Pathways (all corrected p values are <0.0026)

Subjects with severe XPA compared to WT subjects, when exposed to UV, exhibited consistently large effect sizes (0.984) for DNA damage (CPD/6-4 PPs) across all age groups, indicating substantial DNA damage. Large negative effect sizes (-0.984) for DNA NER Core, DNA NER-GG, and DNA NER-TC across all age groups reflect marked deficiencies in DNA repair mechanisms in subjects with severe XPA. Importantly, DNA replication stress also showed large effect sizes (0.984) across all ages, indicating increased replication stress in XPA. ERCC2/XPD displayed large negative effect sizes (-0.984) across all ages, consistent with markedly impaired DNA repair.

#### Genome Instability and Cancer Risk (all corrected p values are <0.0026)

Genome instability (CIN) exhibited large and stable effect sizes (0.984) across all ages, indicating persistent genomic instability in XPA subjects. Skin cancer risks (both melanoma and non-melanoma) generally showed large effect sizes (0.866) across all ages, with a more medium effect size in melanoma risk (0.433) at birth, in keeping with significant overall skin cancer risk.

#### Neurological Outcomes (all corrected p values are <0.0026)

Neuroapoptosis and neurodegeneration showed large effect sizes (0.984 and 0.748 respectively) across all age groups, indicating increased neurological damage in XPA subjects. Neurodevelopmental disorder (NDD) consistently displayed large effect sizes (0.984), reflecting substantial developmental impact. Neuroinflammation showed large effect sizes (0.748) across all ages, indicating elevated inflammation. Neurological symptoms exhibited large effect sizes (0.984), suggesting significant neurological symptoms in subjects with severe XPA exposed to moderate UV exposure. Neuronal oxidative stress showed large effect sizes (0.748) across all ages, indicating increased oxidative stress.

#### Skin Aging and Pigmentation (all corrected p values are <0.0026)

Skin aging, Skin pigmentation and UV photosensitivity consistently displayed large effect sizes (0.984) across all age groups, reflecting significant aging effects, substantial skin pigmentation changes. and marked UV sensitivity.

#### XPA Protein Expression (all corrected p values are <0.0026)

XPA protein expression exhibited large negative effect sizes (-0.984) across all age groups, indicating a marked reduction in XPA expression in XPA subjects compared to WT.

#### Summary

The data reveal substantial differences between WT and severe XPA subjects when both are exposed to moderate UV radiation. Subjects with severe XPA exhibit marked DNA repair deficiencies, increased genomic instability, significant neurological impacts, and heightened skin aging and cancer risk. These findings continue to underscore the critical role of XPA in maintaining genomic stability and highlight the severe consequences of its deficiency.

### (6) Subjects with Moderate XPA versus subjects with severe XPA without UV exposure See Appendix B: Heat Map 6

#### DNA Damage and Repair Pathways (all corrected p values are <0.0026)

Subjects with severe XPA, compared to those with moderate XPA, without UV exposure exhibited somewhat variable but large effect sizes (>0.747) for DNA damage (CPD/6-4 PPs) across all age groups. Large negative effect sizes (-0.984) for DNA NER Core, DNA NER-GG, and DNA NER-TC across all age groups reflect more marked deficiencies in DNA repair mechanisms in severe XPA subjects compared to moderate XPA. DNA replication stress showed effect sizes ranging from small to large (0.197 to 0.984), indicating elevated replication stress with some variation across ages. ERCC2/XPD displayed consistently large and negative effect sizes (-0.984) across all ages, consistent with more impaired DNA repair.

#### Genome Instability and Cancer Risk (all corrected p values are <0.0026)

Genome instability (CIN) exhibited large and stable effect sizes (0.905 to 0.984) across all ages, indicating persistent increased genomic instability in severe XPA subjects. Skin cancer risks (both melanoma and non-melanoma) generally showed large effect sizes (0.748 to 0.984) across all ages, with variations in melanoma risk ranging from 0.669 to 0.827, suggesting significant overall increased skin cancer risk.

#### Neurological Outcomes (all corrected p values are <0.0026)

Neuroapoptosis and neurodegeneration showed large effect sizes (0.748 to 0.984) across all age groups, indicating increased neurological damage in severe XPA subjects compared to moderate XPA. Neurodevelopmental disorder (NDD) consistently displayed large effect sizes (0.984), reflecting substantial developmental impact. Neuroinflammation showed moderate effect sizes (0.748) across all ages, indicating elevated inflammation. Neurological symptoms exhibited large effect sizes (0.984), consistent with significant neurological symptoms in severe XPA. Neuronal oxidative stress also showed large effect sizes (0.748 to 0.787) across all ages, indicating increased oxidative stress.

#### Skin Aging and Pigmentation (all corrected p values are <0.0026)

Skin aging consistently displayed large effect sizes (0.905 to 0.984) across all age groups, reflecting significant aging effects. Skin pigmentation also generally showed variably effect sizes (0.276 to 0.905) across all ages, indicating substantial pigmentation changes.

#### XPA Protein Expression (all corrected p values are <0.0026)

XPA protein expression exhibited large negative effect sizes (-0.984) across all age groups, indicating significantly reduced XPA expression in severe XPA subjects compared to moderate XPA.

#### Summary

The data reveal substantial differences between severe and moderate XPA subjects without UV exposure. Severe XPA subjects exhibit consistently increased DNA repair deficiencies, increased genomic instability, significant neurological impacts, and heightened skin aging and cancer risk. These findings underscore the critical role of XPA in maintaining genomic stability and highlight the severe consequences of its deficiency even in the absence of moderate UV exposure.

### (7) Subjects with Severe XPA versus subjects with severe XPA plus moderate UV exposure See Appendix B: Heat Map 7

#### DNA Damage and Repair Pathways (all corrected p values are <0.0026)

Subjects with severe XPA compared to those with severe XPA exposed to moderate UV radiation exhibited large effect sizes for DNA damage (CPD/6-4 PPs) across all age groups, with modest variability ranging from 0.354 to 0.984. Negative effect sizes (-0.984) for DNA NER Core, DNA NER-GG, and DNA NER-TC across all age groups indicate significant deficiencies in DNA repair mechanisms in severe XPA subjects. DNA replication stress showed variable effect sizes (ranging from -0.590 to 0.905), indicating elevated replication stress with some variable moderate to large decrease at older ages ages. ERCC2/XPD displayed consistently negative effect sizes (-0.905) across all ages, consistent with marked impaired DNA repair.

#### Genome Instability and Cancer Risk (all corrected p values are <0.0026)

Genome instability (CIN) exhibited large and stable effect sizes (0.827 to 0.905) across all ages, indicating persistent genomic instability in severe XPA subjects with moderate UV exposure. Skin cancer risks (both melanoma and non-melanoma) generally showed variably large effect sizes (0.197 to 0.905) across all ages, with variations in melanoma (0.197 to 0.905), suggesting overall significant skin cancer risk.

#### Neurological Outcomes (all corrected p values are <0.0026)

Neuroapoptosis and neurodegeneration showed large effect sizes (0.709 to 0.984) across all age groups, indicating increased neurological damage in severe XPA subjects compared to moderate XPA with UV exposure. Neurodevelopmental disorder (NDD) consistently displayed large effect sizes (0.984), reflecting substantial developmental impact. Neuroinflammation showed large effect sizes (0.709) across all ages, indicating elevated inflammation. Neurological symptoms exhibited large effect sizes (0.984), suggesting a significant difference in neurological symptoms in severe XPA vs moderate XPA. Neuronal oxidative stress showed large effect sizes (0.709) across all ages, indicating increased oxidative stress.

#### Skin Aging and Pigmentation (all corrected p values are <0.0026)

Skin aging consistently displayed large effect sizes (0.905) across all age groups, reflecting significant aging effects. Skin pigmentation generally showed large effect sizes (0.590 to 0.905) across all ages, indicating substantial pigmentation changes. UV photosensitivity consistently also showed large effect sizes (0.984), indicating significant sensitivity.

#### XPA Protein Expression (all corrected p values are <0.0026)

XPA protein expression exhibited negative effect sizes (-0.984) across all age groups, indicating significantly reduced XPA expression in severe XPA subjects compared to those with UV exposure.

#### Summary

The data reveal substantial differences between severe XPA and severe XPA plus UV exposure subjects. Severe XPA subjects exposed to moderate UV radiation exhibit consistently increased DNA repair deficiencies, increased genomic instability, significant neurological impacts, and heightened skin aging and cancer risk compared to those subjects with severe XPA without UV exposure. These findings underscore the critical role of XPA in maintaining genomic stability and highlight the severe consequences of its deficiency, exacerbated by UV radiation.

## Discussion

The present virtual longitudinal study comprehensively evaluates the impact of XPA (xeroderma pigmentosum group A) and UV exposure on DNA damage, genomic stability, neurological outcomes, skin aging, skin cancer and XPA protein expression across six different age groups from birth to age 20 years. Our findings highlight the marked differences between WT (wild type) subjects and subjects with moderate and severe XPA, as well as the exacerbating effects of moderate UV exposure on these conditions.

### DNA Damage and Repair Pathways

Our results demonstrate consistently large effect sizes for DNA damage (CPD/6-4 PPs) across all age groups in both WT and XPA subjects exposed to UV radiation. This aligns with previous studies that have shown UV radiation induces substantial DNA damage, primarily in the form of cyclobutane pyrimidine dimers (CPDs) and 6-4 photoproducts (6-4 PPs) [23]. Notably, DNA repair mechanisms such as NER (nucleotide excision repair) exhibited significant deficiencies in XPA subjects, with negative effect sizes for DNA NER Core, NER-GG (global genomic NER), and NER-TC (transcription-coupled NER). This is consistent with the role of XPA in the NER pathway, which is crucial for repairing UV-induced DNA damage [10,11].

### Genome Instability and Cancer Risk

Large and stable effect sizes for genome instability (CIN) and skin cancer risk across all age groups underscore the persistent risks associated with both XPA and UV exposure. These findings are in line with established research indicating that deficiencies in DNA repair mechanisms significantly increase genomic instability and cancer susceptibility [24]. These data also indicate that moderate UV exposure exacerbates these risks, further highlighting the importance of effective DNA repair pathways in preventing carcinogenesis [11].

### Neurological Outcomes

The deleterious neurological impact of XPA, particularly in younger age groups, is evident from the large effect sizes observed for neurodevelopmental disorder (NDD) and neurological symptoms. This developmental sensitivity to DNA damage has been previously reported, emphasizing the critical role of DNA repair in neurodevelopment [6]. The consistent large effect sizes for neuronal oxidative stress in XPA subjects suggest that oxidative damage may be a contributing factor to the observed neurological deficits [7]. Recent studies have further linked impaired DNA damage response mechanisms to microcephaly and progressive cognitive impairment in individuals with XP [25, 26].

### Skin Aging and Pigmentation

The large effect sizes for skin aging and pigmentation changes in XPA subjects, both with and without UV exposure, are consistent with the clinical manifestations of xeroderma pigmentosum. The increased photosensitivity in younger individuals further underscores the need for protective measures against UV radiation, particularly in susceptible populations [1,3,8].

### XPA Protein Expression

The observed negative effect sizes for XPA protein expression across all age groups in XPA subjects compared to WT highlight the significant deficiency in XPA expression in these individuals. This aligns with the known loss of function (LOF) mutations in XPA patients that lead to reduced (moderate XPA) or non-functional XPA protein (severe XPA), compromising the NER pathway and increasing susceptibility to DNA damage and its associated risks [11].

### Comparisons and Implications

The comparison between moderate and severe XPA subjects reveals a gradient of severity, with severe XPA subjects exhibiting more pronounced deficiencies in DNA repair and higher risks of genomic instability, skin cancers and neurodevelopmental delays (NDD). The addition of UV exposure further exacerbates these differences, emphasizing the combined effects of environmental factors on genetic conditions [10,24].

### Conclusions and Future Directions

The findings from this study provide important insights into the mechanisms underlying XPA and the exacerbating effects of UV exposure. These results underscore the importance of early detection and intervention in managing XPA and highlight the need for rigorous UV protection strategies, particularly in younger individuals and those with compromised DNA repair mechanisms. Future research should focus on exploring therapeutic approaches including AI assisted virtual drug trials [13,14] to enhance DNA repair capacity in XPA and other XP patients and further investigating the long-term neurological impacts of DNA repair deficiencies.

A detailed assessment of the Advantages and Limitations of this aiHumanoid based virtual longitudinal study of XPA is presented in Appendix C.

## Data Availability

All data produced in the present study are available upon reasonable request to the author.

## Acknowledgement

The author wants to thank Arjen F. Thiel PhD of Erasmus University Medical Centre, Rotterdam, the Netherlands for his expert insights and estimates of residual protein expression and function in XPA and other XP complementation groups. Dr. Thiel’s work is focused on dissecting the molecular mechanisms and regulation of DNA repair pathway nucleotide excision repair (NER). In addition, he studies the biological consequences of pathogenic mutations in NER and NER-related genes and their impact on health and disease by cell biological analysis of patient-derived cell lines.

## Appendix A: reference for 19 XP/XPA features from the published literature

<u>DNA Damage and Repair Features</u>: (1) DNA_Damage_CPD/6-4_PPs, (2) DNA_NER_Core, (3) DNA_NER-GG, (4) DNA_NER-TG, (5) DNA_Replication Stress and (6) Genome Instability

Neurologic Features: (7) Neuroapoptosis, (8) Neurodegeneration, (9)NeuroDevelopmental Disorder/NDD, (10) Neuroinflammation, (11) Neurological Symptoms, (12) Neuronal Oxidative Stress

<u>Skin Features</u>: (13) Skin-Ageing, (14) Skin-Pigmentation, (15) Melanoma, (16) Nonmelanoma skin cancer, (17) UV_Photosensitivity

<u>XP Protein Expression</u>: (18) XPA expression, (19) ERCC2/XPD

## Appendix B: Heat Maps for the 7 different Genotypic Cohorts Representing the Age vs XP Feature Table for Cliff’s delta (d) Effect Size Estimates

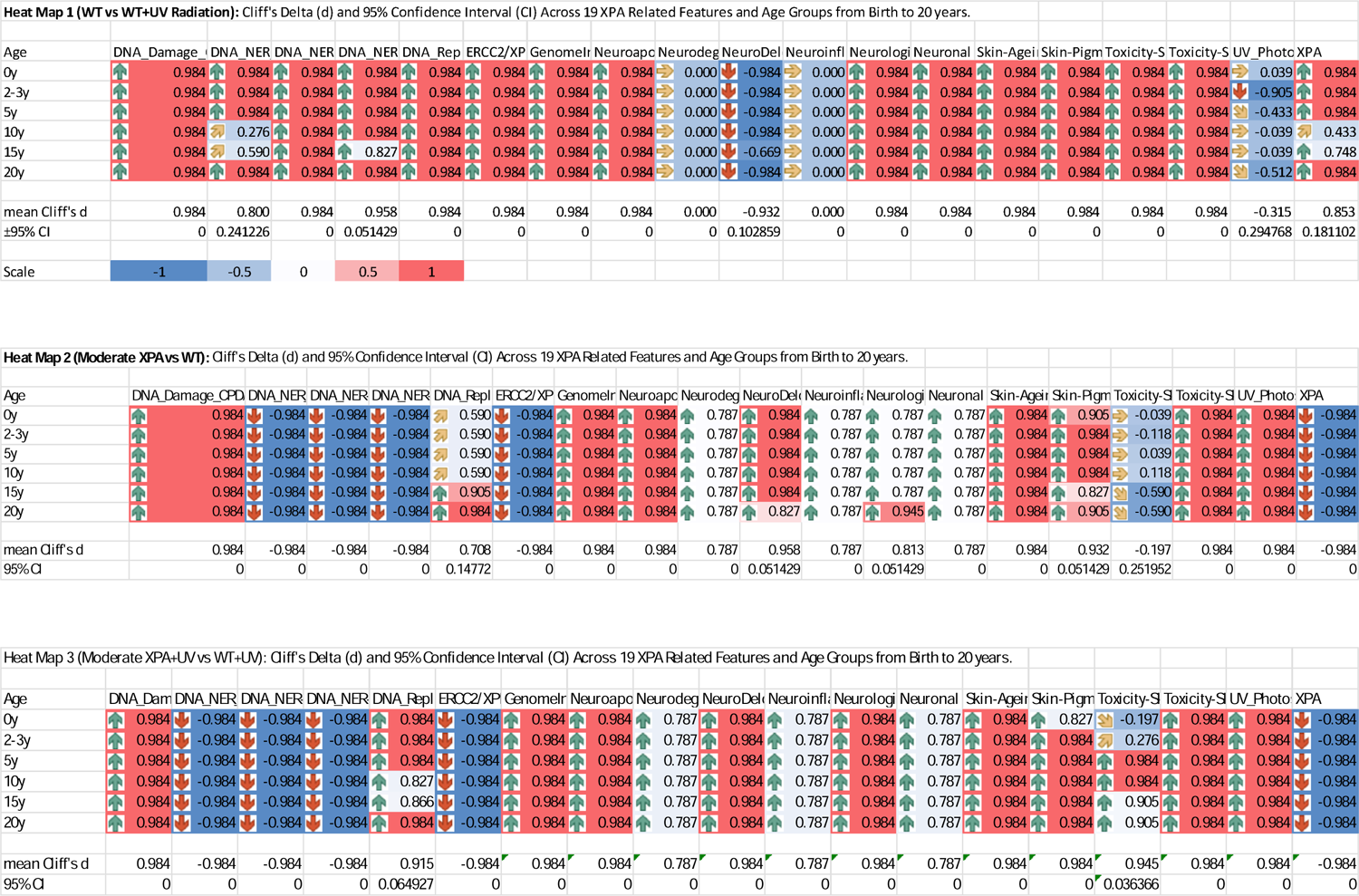
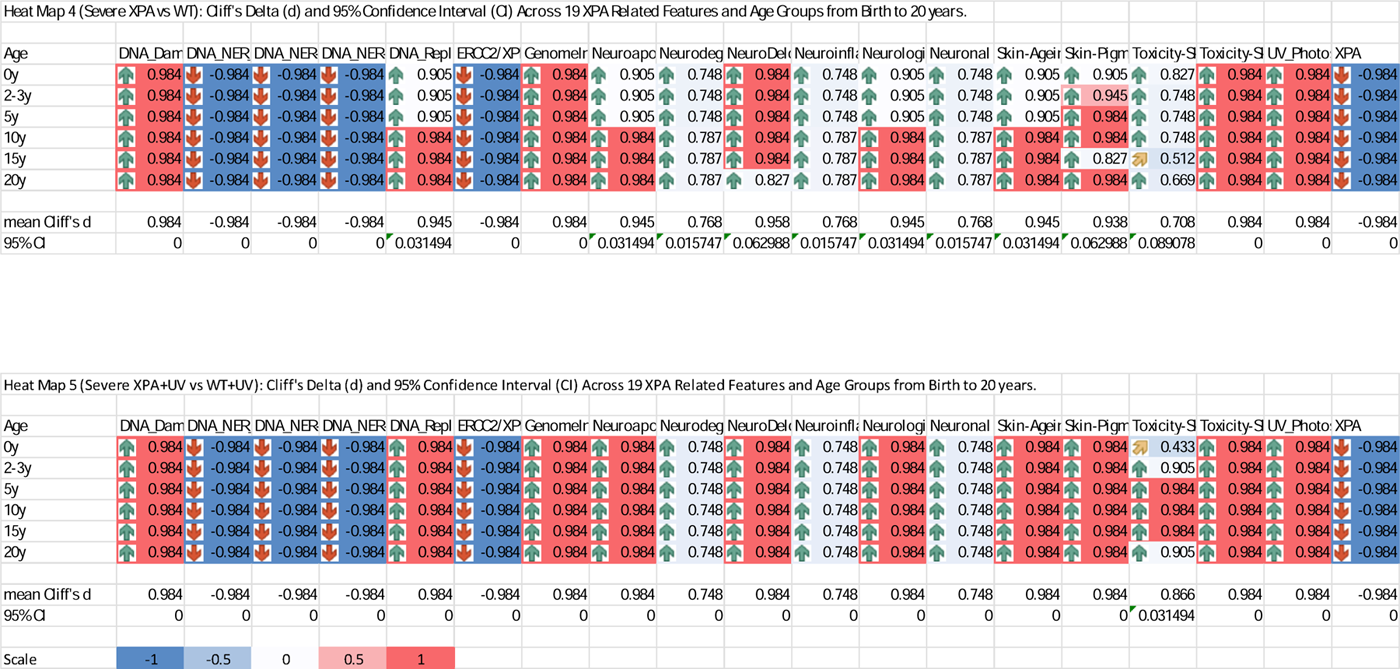
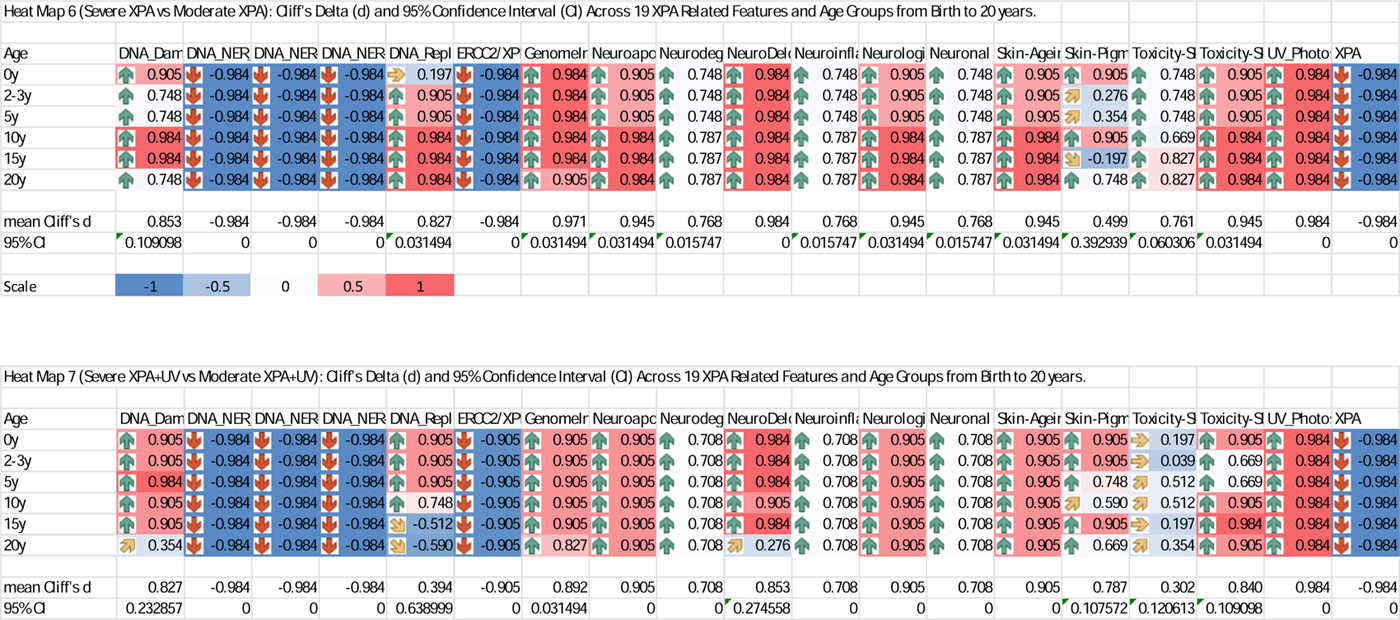

## Appendix C: Analysis of Advantages, Limitations, and Ethical Considerations Associated with this AI-Assisted Virtual Longitudinal Study of XPA in Children and Adolescence

### Advantages

Advanced AI Technology: The aiHumanoid platform offers an innovative method for simulating XPA progression, minimizing ethical concerns and expanding research capabilities for rare genetic disorders.

Comprehensive Analysis: This virtual study provides a detailed evaluation of the effects of UV exposure on DNA repair mechanisms, cancer risks, and neurological outcomes in individuals with XPA, delivering profound insights into the disorder’s complex impact.

Controlled Environmental Simulation: Simulated moderate UV exposure scenarios enable precise studies of environmental effects on disease progression.

Efficient Longitudinal Design: Observes disease markers over time, circumventing the logistical and financial challenges associated with traditional long-term patient follow-up.

Complete Dataset: Lack of participant attrition ensures data integrity and significantly boosts the reliability of longitudinal analysis.

Rigorous Statistical Methodology: Employs advanced statistical techniques, including the Wilcoxon sign rank test and Cliff’s delta, for accurate data evaluation.

### Limitations

Model Reliance: The findings heavily depend on simulation accuracy, which currently is based on data covering approximately 35% of the human genome, potentially limits the assessment of genetic interactions.

Generalizability: Findings from virtual subjects may not entirely reflect real-world population dynamics, impacted by biological and environmental variability.

Limited Age Focus: The study concentrates on individuals from birth to age 20, and does not capture, by design, the long-term outcomes of XPA, which are essential for a more comprehensive understanding of the disorder.

Risk of Overestimation: Controlled simulation conditions might not fully represent the complexities of real-life environments, possibly leading to overestimated effects.

Simplistic UV Exposure Model: The UV exposure simulation may not adequately reflect the variability and intensity of real-world UV exposure, potentially affecting the study’s applicability to actual environmental conditions.

### Future Directions

The ongoing development of aiHumanoid simulations aims to cover approximately 99% of the human genome over the next 3-5 years, increasingly relying on real-world wet lab and early-stage clinical trial outcomes.

